# The Canadian multi-ethnic research on aging (CAMERA) study: Study design, participant characteristics, and preliminary findings

**DOI:** 10.1101/2025.10.31.25338992

**Authors:** Tulip Marawi, Harleen Rai, Rohina Kumar, Katie L. Vandeloo, Rachel Yep, Madeline Wood Alexander, Silina Z. Boshmaf, Sarah-Mei Chen, Georgia Gopinath, Simran Malhotra, Angelina Zhang, Alexander J. Nyman, Lucas Xavier Perri, Valery Sit, Tallinn F. L. Splinter, Douglas P. Munoz, Walter Swardfager, Jennifer D. Ryan, Sandra E. Black, Maged Goubran, Jennifer S. Rabin

**Author notes:** **Corresponding Author**: Jennifer S. Rabin, Hurvitz Brain Sciences Program, Sunnybrook Health Sciences Centre, Room M6-178, 2075 Bayview Avenue, Toronto, ON M4N 3M5, Canada. Co-senior authors (Maged Goubran, Jennifer S. Rabin).

## Abstract

**INTRODUCTION:** South Asian and Chinese individuals are the largest and fastest-growing ethnoracial groups in Canada, yet they remain underrepresented in dementia research. To address this gap, we established the <u>CA</u>nadian <u>M</u>ulti-<u>E</u>thnic <u>R</u>esearch on <u>A</u>ging (CAMERA) study.

**METHODS:** CAMERA is a longitudinal observational study conducted in Toronto, Canada, enrolling 300 adults aged 55–85 who self-identify as South Asian, Chinese, or non-Hispanic White (NHW). Participants complete in-person visits at baseline, Year 3, and Year 5, which include clinical and cognitive assessments, brain MRI, and blood biomarkers. Annual remote questionnaires track health and lifestyle.

**RESULTS:** Among 200 participants, vascular and metabolic profiles differed across groups. South Asian and Chinese participants reported greater cognitive concerns than NHW participants and had lower MoCA scores, driven primarily by language-heavy and culturally dependent items. Eye-tracking measures did not differ across groups.

**DISCUSSION:** CAMERA provides a deep phenotyping framework to investigate dementia risk and resilience in Asian Canadians.

## 1.0 Background

Alzheimer’s disease (AD) and related dementias (ADRD) represent major and growing public health challenges, with substantial personal, societal, and economic impacts.^1^ Despite their increasing burden, much of the existing research in North America has been conducted in predominantly non-Hispanic White (NHW) cohorts. This focus has limited our understanding of how ADRD risk and resilience differ across racial and ethnic populations,^2–6^ which is critical for developing effective and equitable prevention and intervention strategies.

This knowledge gap is especially pronounced among individuals of Asian descent.^2,7–9^ In Canada, studies of aging and ADRD report markedly low levels of Asian representation^10,11^ relative to their population share (approximately 20%).^12^ Comparable patterns of underrepresentation are evident in the U.S. For example, in the National Alzheimer’s Coordinating Center (NACC) Uniform Data Set, only 2.6% of participants are of Asian descent, despite Asian individuals comprising nearly 14% of older adults in the U.S.^13,14^

Beyond underrepresentation, a major limitation of the existing literature is the routine aggregation of heterogeneous Asian subpopulations into a single, monolithic “Asian” category.^15^ This limitation is especially consequential for South Asian and Chinese populations, which are the two largest and fastest-growing Asian groups in Canada.^12^ Treating these groups as homogeneous obscures meaningful between-group differences across a range of clinically relevant factors, including vascular risk burden, a well-established contributor to cognitive impairment and dementia.^16,17^

Evidence from Canadian and U.S. studies consistently demonstrates substantial heterogeneity in vascular risk profiles across Asian subpopulations.^18–20^ Specifically, South Asian individuals tend to exhibit a higher vascular risk burden than both Chinese and NHW individuals, with this elevated risk often emerging at younger ages. In contrast, Chinese individuals generally have a lower vascular risk burden than these groups.^18–22^ However, some evidence also suggests that Chinese immigrant populations have a higher vascular risk burden compared to individuals living in China, with risk increasing with longer duration of residence in the host country.^23^ Although epidemiological data on dementia incidence among Asian subgroups remain limited, available evidence aligns with these patterns. For example, U.S. data indicate that South Asian individuals may have a higher incidence of dementia than Chinese individuals and some other ethnoracial groups;^24^ these differences appear to be particularly pronounced among individuals with diabetes and less evident among those without diabetes.^25^

Given this heterogeneity, there is a clear need for longitudinal research to characterize ADRD risk and resilience in Asian subgroups. Subgroup-specific insights are critical, as differences in vascular burden, metabolic profiles, and sociocultural factors may influence disease trajectories and inform prevention and intervention efforts. Because ADRDs develop gradually over decades, with neuropathology emerging long before symptoms appear, biomarker-informed longitudinal cohorts provide a powerful means to detect early biological changes and evaluate whether disease pathways differ across ethnoracial groups.

Canada is particularly well positioned to support this work. South Asian (i.e., Indian, Pakistani, Sri Lankan, Bangladeshi) and Chinese communities together comprise a substantial and growing proportion of the Canadian population, accounting for 7.1% and 4.7% of Canadians, respectively.^12^ In major urban centers, such as Toronto, these proportions are even higher, with South Asian and Chinese communities comprising 14% and 10.7% of the population.^12^ This demographic context provides an important opportunity to conduct longitudinal, biomarker-informed research in groups that have been historically underrepresented in ADRD studies.

To leverage this opportunity, we established the CAnadian Multi-Ethnic Research on Aging (CAMERA) study, a longitudinal observational cohort based in Toronto that investigates early mechanisms underlying ADRD risk and resilience among underrepresented South Asian and Chinese populations. CAMERA employs a deep phenotyping approach, with participants completing comprehensive biannual assessments, including clinical evaluations, blood-based biomarker assessments, questionnaires, and both standard and novel cognitive tests designed to minimize language demands and cultural influences. By integrating biomarker, clinical, and cognitive data, CAMERA aims to advance understanding of population-specific biological pathways and inform future ADRD prevention and intervention strategies.

## 2.0 Methods

The CAMERA study was approved by the Research Ethics Board at Sunnybrook Health Sciences Centre in Toronto, Canada.

### 2.1 Participants

The CAMERA study is an ongoing longitudinal cohort that aims to recruit and retain 300 community-dwelling adults aged 55 to 85 years who self-identify as South Asian, Chinese, or NHW (100 participants per group).

According to recent census data, approximately 71% and 56% of South Asian and Chinese adults over the age of 65 can converse in English.^26^ Given this level of language proficiency along with the practical and resource considerations involved in validating and administering assessments in multiple languages, all study assessments are conducted in English. However, to support participants and create a welcoming testing environment, language assistance is available in Punjabi, Hindi, Urdu, Cantonese, and Mandarin.

Eligibility criteria for the study include: (1) self-identification as South Asian, Chinese, or NHW; (2) sufficient English proficiency to provide informed consent and complete study assessments. This was assessed at screening using a 5-point Likert scale (1 = none, 3 = adequate, 5 = proficient) for speaking, understanding, and reading English. Participants are required to score ≥3 in all domains, with proficiency additionally confirmed by the study screener; (3) a minimum of 8 years of formal education to support meaningful participation in study assessments and questionnaires, which are written at a grade 6–8 reading level; (4) a global Clinical Dementia Rating (CDR)^27^ of 0 (clinically normal) or 0.5 (mild cognitive difficulties) at baseline; (5) Adequate (or corrected) vision and hearing to participate in cognitive testing; and (6) availability of a study partner who knows the participant well, maintains at least weekly contact, and can provide informed observations about their cognition and function.

Exclusion criteria were designed to minimize confounding factors and to exclude conditions that could obscure associations of interest, including: (1) history of major cardio/cerebrovascular event (e.g., myocardial infarction, symptomatic stroke); (2) presence of unstable medical conditions (e.g., pulmonary or endocrine disorders); (3) active infectious diseases; (4) current diagnosis of major psychiatric disorders, neurological disorders, or dementia; (5) history of a significant learning disability; (6) history of a significant head trauma or recurrent concussions with loss of consciousness or hospitalization; (7) pain or sleep disorders that could interfere with cognitive testing; (8) recent history of substance or drug abuse; (9) medical conditions that may obscure relevant relationships (e.g., active cancer); (10) MRI contraindications; and (11) current enrollment in an intervention study.

### 2.2 Study Design

Participants complete in-person assessments at baseline (Year 1), Year 3 and Year 5. Each in-person assessment involves two in-person study visits scheduled within a 1-month window. These visits involve comprehensive clinical assessments (vital signs, anthropometric measures, gait assessment, hearing test), brain MRI, a blood draw, cognitive testing (including eye tracking tasks), as well as questionnaires. Questionnaires can be completed on-site or remotely via REDCap. At the end of the first visit, participants are provided with an actigraphy device to wear for 12 consecutive days to monitor physical activity and sleep patterns. During the intervening years (Years 2 and 4), participants complete questionnaires remotely via REDCap (**Figure 1**).

**Figure 1.**
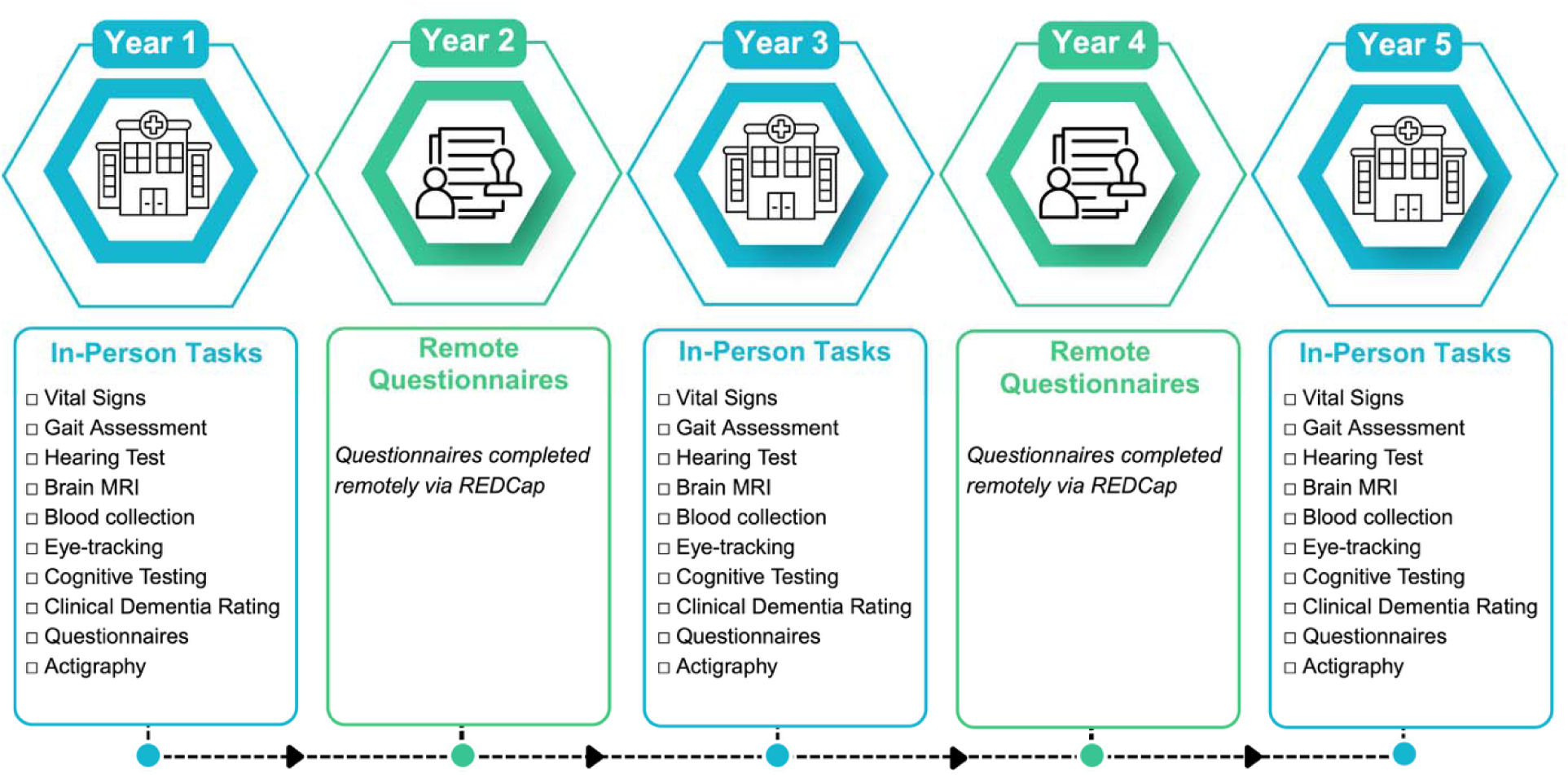
Timeline and assessments. The figure depicts the study timeline and procedures of the CAMERA study across the five-year study period. Participants complete in-person assessments at baseline (Year 1), Year 3 and Year 5, and complete questionnaires annually on REDCap.

### 2.3 Study Measures

#### 2.3.1 Clinical assessments

##### Clinical Dementia Rating (CDR)

Participants and their study partners complete the CDR,^27^ which is administered by trained personnel at baseline (Year 1), Year 3, and Year 5.

##### Vital Signs

Blood pressure and heart rate are measured using an automated sphygmomanometer. Six readings (three seated and three standing) are taken one minute apart. Respiration rate, waist circumference, hip circumference, height, and weight are also collected.

##### Gait Assessment

Gait is measured using a pressure-sensitive walkway (GaitRite)^28^ that records the timing and placement of each step using protocols from the Ontario Neurodegenerative Disease Research Initiative (ONDRI).^10,29^ Gait performance is assessed under both single-task (preferred and fast walking) and cognitively demanding dual-task conditions. There are three dual-tasks: (1) counting backwards from 100 by ones; (2) counting backwards from 100 by sevens; and (3) naming as many different animals as possible. These cognitive tasks are included solely to create a dual-task condition and are not scored. All gait assessments take place after neuropsychological testing to avoid practice effects with similar tests.

##### Hearing Test

Participants complete a hearing assessment using the Triple Digit Test.^30,31^ In this task, participants listen to a voice recording in which three digits are presented against different levels of background noise and report the digits in the correct order.

#### 2.3.2 Brain Magnetic Resonance Imaging (MRI)

Brain MRI scans are acquired on a 3T Siemens MAGNETOM Prisma magnetic resonance system, using a 64-channel head coil. Each 50-minute scanning session includes the following sequences (see **Supplementary Table 1** for more details): 1) 3-dimensional T1-weighted magnetization prepared rapid gradient echo (MPRAGE) acquired in the sagittal plane; 2) 3-dimensional T2-weighted Sampling Perfection with Application optimized Contrast using different flip angle Evolution (SPACE) acquired in the axial plane; 3) 3-dimensional T2-weighted Fluid-Attenuated Inversion Recovery (FLAIR) acquired in the sagittal plane; 4) resting-state functional MRI (rs-fMRI) acquired in two phase-encoding (PE) directions (anterior-to-posterior, and posterior-to-anterior); 5) multi-shell diffusion weighted imaging (DWI) acquired with 95 diffusion directions in two PE directions (anterior to posterior (b=2000s/mm^2^, b=1000s/mm^2^) and posterior-to-anterior (b=0s/mm^2^) for distortion correction); 6) a high-resolution T2-weighted turbo spin echo (TSE) hippocampal scan, oblique to the long axis of the hippocampus; 7) pseudo-continuous arterial spin labeling (ASL); and 8) susceptibility weighted imaging (SWI).

#### 2.3.3 Cognitive Assessments

##### Eye tracking

A major barrier to conducting ADRD research in diverse ethnoracial groups is that most cognitive tests were developed and validated in English-speaking NHW individuals born in Anglosphere countries.^32^ As a result, their validity may be compromised when administered to adults from different linguistic and cultural backgrounds.^32,33^ Strategies to address this limitation include translating existing tests, developing group-specific norms, and/or using an interpreter to conduct neuropsychological testing.^34,35^ However, these approaches are resource-intensive and may not fully eliminate bias. An alternative approach employed in the CAMERA study is to incorporate tests that minimize language demands inherent to task performance (beyond the instructions themselves) and reduce cultural influences. As part of this approach, we include eye-tracking tasks, a widely accepted method for assessing cognition that has been applied in non-human primates,^36–38^ infants,^39–41^ and adults with aphasia or speech limitations.^42,43^ Eye-tracking tasks are particularly advantageous because they can be designed to minimize language-dependent and culturally biased stimuli and do not require verbal responses, making them especially well-suited for participants from diverse linguistic backgrounds.

The CAMERA study employs two well-validated eye-tracking tasks: the Interleaved Pro/Anti-Saccade Task (IPAST)^44,45^ and the Visual Paired Comparison Task (VPCT).^46^ The IPAST assesses processing speed and executive function, whereas the VPCT assesses episodic memory.

In the IPAST, participants complete intermixed pro- and anti-saccade trials, in which they are instructed either to look toward a peripheral visual stimulus (pro-saccade) or to inhibit this response and look in the opposite direction (anti-saccade)^44,45^ (see **Figure 2A**). Saccadic reaction time on pro-saccade trials provides a measure of processing speed through sensorimotor pathways, whereas saccadic reaction time and direction errors on anti-saccade trials provide measures of executive function.^44,45,47^ Previous studies demonstrate that both pro- and anti-saccade performance declines with age^48–50^, and that anti-saccade performance is impaired in mild cognitive impairment and dementia.^51,52^

**Figure 2.**
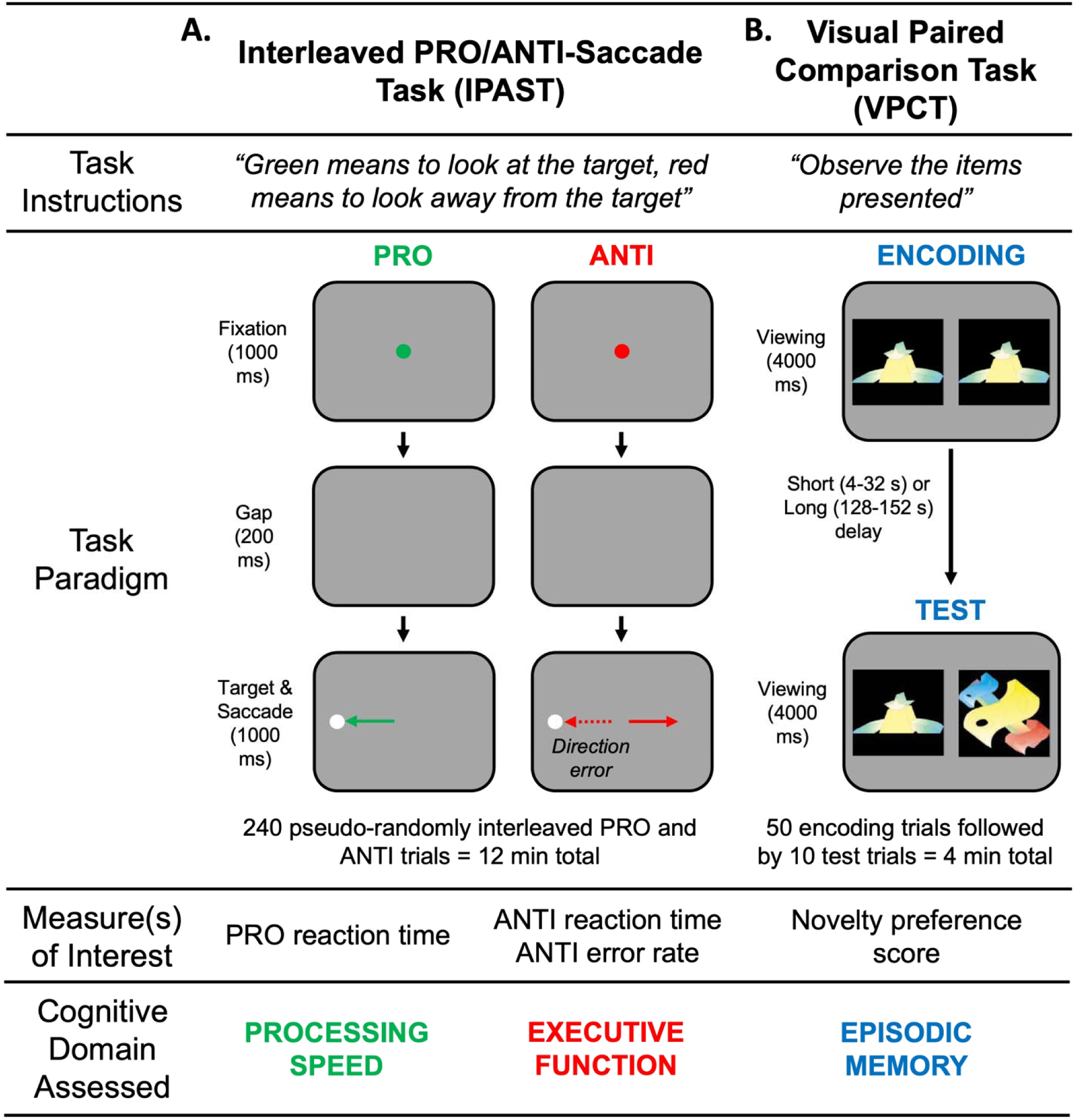
Eye tracking measures. **A**) depicts the Interleaved PRO/ANTI-Saccade Task (IPAST) and **B)** depicts the Visual Paired Comparison Task (VPCT).

The VPCT begins with a familiarization phase in which participants are presented with pairs of identical abstract images^46^ (see **Figure 2B**). In the test phase, participants are presented with a new series of image pairs, each consisting of one “old” image from the familiarization phase and one “novel” image not previously seen.^46^ Based on the well-established novelty effect,^53,54^ participants with intact memory are expected to spend more time viewing the novel image than the old image (i.e., novelty preference). Prior work shows that novelty preference on the VPCT declines with age,^55^ is impaired in mild cognitive impairment,^53^ and predicts progression to AD dementia.^56,57^

##### Standard Neuropsychological Tests

In addition to eye-tracking tasks, participants complete a battery of standardized neuropsychological tests assessing processing speed, executive function, episodic memory, language, and visuospatial abilities (see **Table 1**). The battery includes both paper- and computer-based assessments and was designed to minimize language demands and culturally specific stimuli. However, a subset of commonly used language-based measures is included to facilitate comparison with existing literature, despite the potential for cultural and linguistic bias in these assessments.

**Table 1.** CAMERA Neuropsychological Assessment Battery.

| **Cognitive Domain** | **Tests** |
| --- | --- |
| **Global Cognition** | Montreal Cognitive Assessment (MoCA)<br>Clinical Dementia Rating (CDR) Scale |
| **Processing Speed** | Digit Symbol-Coding subtest from the Wechsler Adult Intelligence Scale (Third Edition)<br>Trails Making Test (TMT) Part A<br>Color Trails Test (CTT) I |
| **Executive Function** | N-Back Task (1-Back and 2-Back)<br>Sustained Attention to Response Task (SART)<br>Trails Making Test (TMT) Part B<br>Color Trails Test (CTT) II<br>Phonemic Fluency (FAS) |
| **Visuoperceptual/**<br>**Visuospatial**<br>**Processing** | Four Choice Oddity Task<br>Benson Complex Figure Test – Copy Trial |
| **Learning and**<br>**Memory** | Free and Cued Selective Reminding Test (Form A)<br>7/24 Learning, Immediate Recall, and Delayed Recall (Form I)<br>Benson Complex Figure Test – Delayed |
| **Language** | Category Fluency (Animals, Fruits, and Vegetables) |
| **Pattern**<br>**Separation** | Doors and People Test (Doors Subtest)<br>Mnemonic Similarity Task (MST – Set C) |

#### 2.3.4 Questionnaires

Participants complete a series of questionnaires annually via REDCap (see **Supplementary Table 2**). The questionnaires capture a range of variables, including: 1) demographic information; 2) socioeconomic status; 3) immigration history; 4) linguistic background; 5) acculturation; 6) environmental and community factors; 7) medical history; 8) mental health and personality characteristics; 9) physical activity and sedentary behavior; 10) sleep quality; 11) quality of life; 12) perceived discrimination; 13) cognitive concerns; 15) functional status; and 16) sex-specific health history.

#### 2.3.5 Actigraphy

Participants are provided with a wearable wrist accelerometer (GENEActive) that continuously records daytime and nighttime activity. Participants are asked to wear the accelerometer for 12 consecutive days.^58^

#### 2.3.6 Blood Collection

Participants undergo a fasting venous blood draw in the morning. Blood samples are processed at Sunnybrook Health Sciences Centre, and analyzed to measure lipids (i.e., total cholesterol, low-density lipoprotein, high-density lipoprotein, and triglycerides), glucose regulation (i.e., hemoglobin A1c, HbA1c), and inflammation (i.e., C-reactive protein). Serum, plasma, and white blood cell samples are also stored for future analyses, including apolipoprotein E (*APOE*) ε4 genotyping, as well as markers of AD and neurodegeneration, and more detailed inflammatory profiling.

### 2.4 Community-Based Participatory Research Approaches

The Community Advisory Board (CAB) for the CAMERA study was established in January 2024 to ensure that study logistics, including implementation, assessments, and dissemination, are guided by community perspectives. Members were recruited through partnerships with local community organizations, outreach events, and word-of-mouth referrals. The CAB currently includes six community representatives whose identities and lived experiences reflect those of the CAMERA cohort, including two South Asian members, three Chinese members, one NHW members (mean age = 61.3 years, mean years of education = 19 years). Most members also have lived experience with dementia as care partners. Several members later chose to enroll as study participants, providing firsthand experience with study procedures and enhancing the relevance of their feedback.

The CAB meets quarterly, with additional ad hoc meetings as needed, providing ongoing input across the study lifecycle. This includes aligning research objectives with community priorities, supporting community outreach and recruitment, reviewing study materials for cultural appropriateness, and co-developing accessible strategies for knowledge dissemination.

### 2.5 Participation Benefits

A core value of the CAMERA study is a strong commitment to sharing individualized results with participants. Sharing results promotes transparency, builds trust, and acknowledges participants as valued contributors to the research.^59,60^ Participants receive feedback in four domains: (1) <u>MRI results:</u> Participants may request access to their raw MRI data and radiology report. Regardless of whether access to results is requested, incidental findings are communicated to participants, and referrals are arranged within the hospital when feasible. (2) <u>Cognitive testing:</u> Participants may request a feedback session with a neuropsychologist to review their cognitive results. If testing identifies previously unrecognized cognitive impairment, participants are informed and referred to a specialist. (3) <u>Blood test results:</u> Participants may request results from the standard blood panel (e.g., lipid profile and glucose measures).

Clinically relevant findings are reviewed with the participant, and with consent, shared with the participant’s primary care physician. (4) <u>Mental health results</u>. When questionnaire responses indicate elevated symptoms of depression or anxiety, participants are informed and encouraged to discuss these findings with their primary care provider. Participants are also offered relevant resources, including information on free or low-cost, culturally appropriate counselling services.

### 2.6 Recruitment and Retention Strategies

Recruitment focuses on culturally tailored outreach within South Asian and Chinese communities in the Greater Toronto Area. This includes community-based presentations on dementia risk factors, where the CAMERA study is introduced and attendees are invited to participate. Additional strategies include targeted advertising in senior and community centres, newsletters and websites serving South Asian and Chinese communities, and outreach through social media. Word-of-mouth referrals have also greatly supported enrollment, reflecting participants’ positive experiences with the study.

To support retention and foster a sense of engagement among participants, we emphasize participant-centered practices throughout the study. These include sharing individualized results, offering follow-up support and referrals when incidental findings arise, and providing compensation for participants’ time and effort. To maintain ongoing connection with participants, we send quarterly newsletters that include study updates, highlights from recent research, and educational content related to brain health.

## 3.0 Results

### 3.1 Participants Characteristics

Recruitment for the CAMERA study began in mid 2022 and is ongoing. The current analysis includes participants enrolled through July 2025. Baseline demographic and clinical characteristics are summarized in **Table 2**. As of July 2025, 200 participants had completed baseline (Year 1) assessments, including 52 South Asian participants (42 Indian, 3 Pakistani, 2 Bangladeshi, and 5 unspecified), 77 Chinese participants, and 71 NHW participants. Among Chinese participants, self-identified subgroups included 19 Chinese (e.g., Zhuang, Hui, Manchu), 17 Han Chinese, and 24 Hong Kongers. An additional 10 participants selected more than one subgroup (4 Chinese and Han Chinese; 3 Han Chinese and Hong Konger; 2 Chinese and Hong Konger; 1 selected all three), and 7 did not specify a subgroup.

**Table 2.**
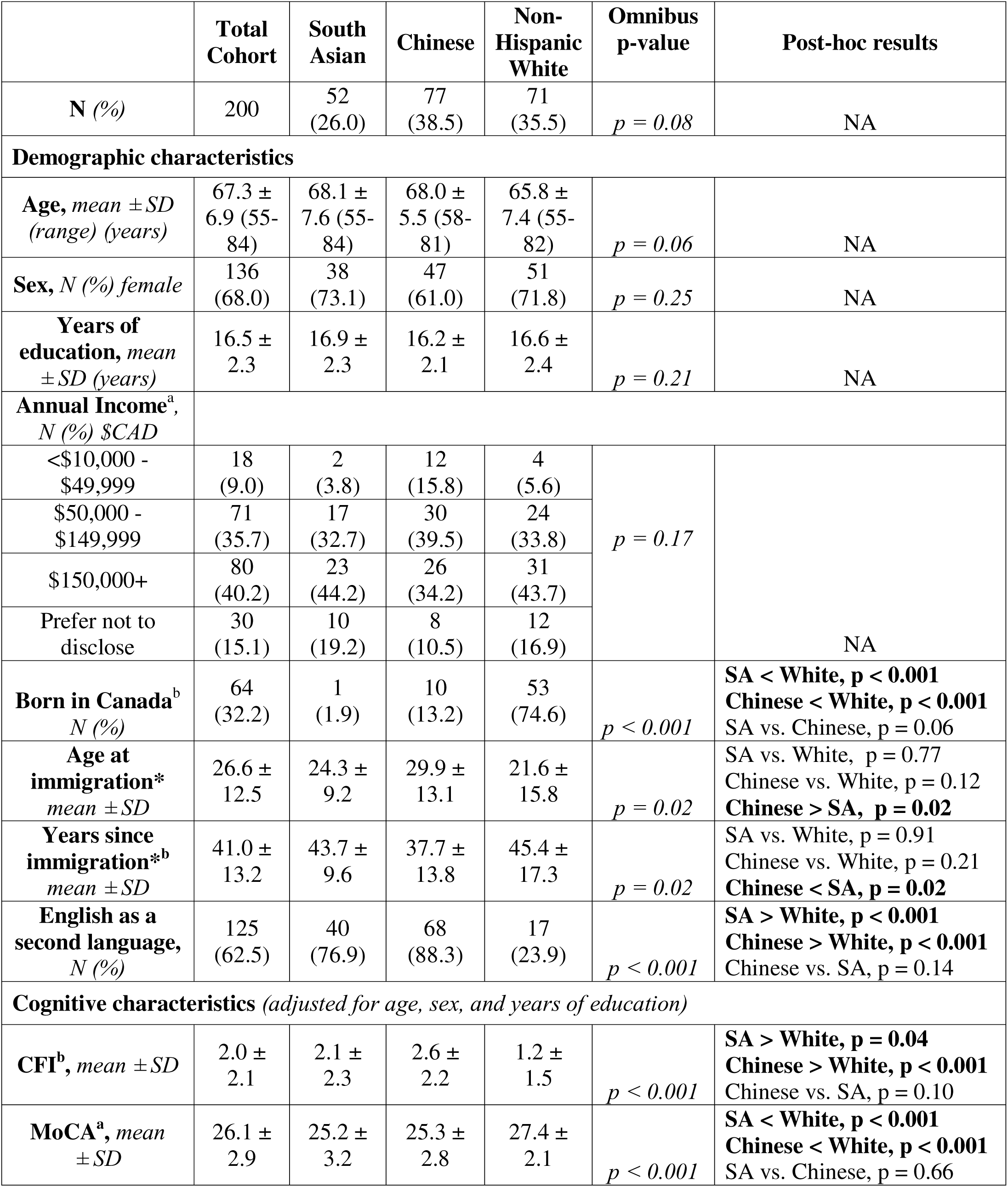
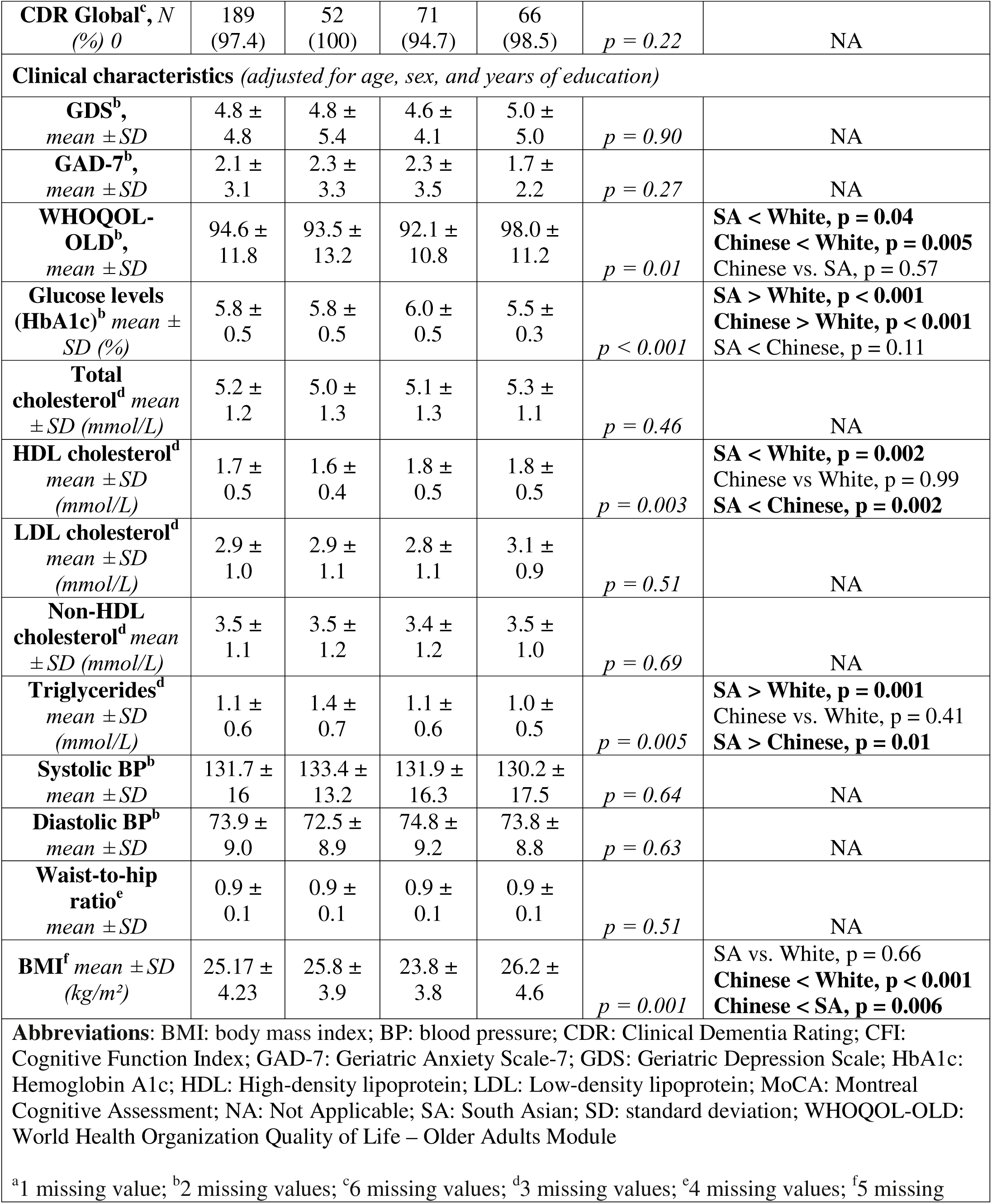
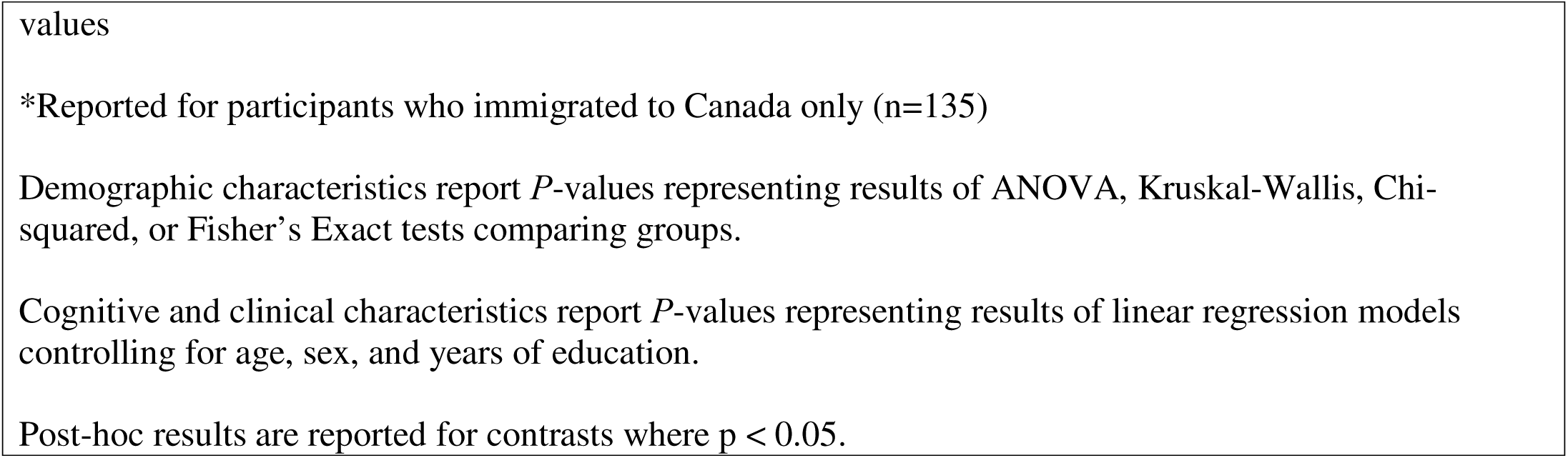
Participant Demographic, Cognitive, and Clinical Characteristics at Baseline (Year 1)

|  | **Total Cohort** | **South Asian** | **Chinese** | **Non-Hispanic White** | **Omnibus p-value** | **Post-hoc results** |
| --- | --- | --- | --- | --- | --- | --- |
| **N (%)** | 200 | 52 (26.0) | 77 (38.5) | 71 (35.5) | $p = 0.08$ | NA |
| **Demographic characteristics** |  |  |  |  |  |  |
| **Age, mean ± SD (range) (years)** | 67.3 ± 6.9 (55-84) | 68.1 ± 7.6 (55-84) | 68.0 ± 5.5 (58-81) | 65.8 ± 7.4 (55-82) | $p = 0.06$ | NA |
| **Sex, N (%) female** | 136 (68.0) | 38 (73.1) | 47 (61.0) | 51 (71.8) | $p = 0.25$ | NA |
| **Years of education, mean ± SD (years)** | 16.5 ± 2.3 | 16.9 ± 2.3 | 16.2 ± 2.1 | 16.6 ± 2.4 | $p = 0.21$ | NA |
| **Annual Income<sup>a</sup>, N (%) \$CAD** | | | | | | |
| <\$10,000 - \$49,999 | 18 (9.0) | 2 (3.8) | 12 (15.8) | 4 (5.6) | $p = 0.17$ | NA |
| \$50,000 - \$149,999 | 71 (35.7) | 17 (32.7) | 30 (39.5) | 24 (33.8) | | |
| \$150,000+ | 80 (40.2) | 23 (44.2) | 26 (34.2) | 31 (43.7) | | |
| Prefer not to disclose | 30 (15.1) | 10 (19.2) | 8 (10.5) | 12 (16.9) |  |  |
| **Born in Canada<sup>b</sup> N (%)** | 64 (32.2) | 1 (1.9) | 10 (13.2) | 53 (74.6) | $p < 0.001$ | **SA < White, $p < 0.001$**<br>**Chinese < White, $p < 0.001$**<br>SA vs. Chinese, $p = 0.06$ |
| **Age at immigration\* mean ± SD** | 26.6 ± 12.5 | 24.3 ± 9.2 | 29.9 ± 13.1 | 21.6 ± 15.8 | $p = 0.02$ | SA vs. White, $p = 0.77$<br>Chinese vs. White, $p = 0.12$<br>**Chinese > SA, $p = 0.02$** |
| **Years since immigration\*<sup>b</sup> mean ± SD** | 41.0 ± 13.2 | 43.7 ± 9.6 | 37.7 ± 13.8 | 45.4 ± 17.3 | $p = 0.02$ | SA vs. White, $p = 0.91$<br>Chinese vs. White, $p = 0.21$<br>**Chinese < SA, $p = 0.02$** |
| **English as a second language, N (%)** | 125 (62.5) | 40 (76.9) | 68 (88.3) | 17 (23.9) | $p < 0.001$ | **SA > White, $p < 0.001$**<br>**Chinese > White, $p < 0.001$**<br>Chinese vs. SA, $p = 0.14$ |
| **Cognitive characteristics (adjusted for age, sex, and years of education)** |  |  |  |  |  |  |
| **CFI<sup>b</sup>, mean ± SD** | 2.0 ± 2.1 | 2.1 ± 2.3 | 2.6 ± 2.2 | 1.2 ± 1.5 | $p < 0.001$ | **SA > White, $p = 0.04$**<br>**Chinese > White, $p < 0.001$**<br>Chinese vs. SA, $p = 0.10$ |
| **MoCA<sup>a</sup>, mean ± SD** | 26.1 ± 2.9 | 25.2 ± 3.2 | 25.3 ± 2.8 | 27.4 ± 2.1 | $p < 0.001$ | **SA < White, $p < 0.001$**<br>**Chinese < White, $p < 0.001$**<br>SA vs. Chinese, $p = 0.66$ |
| **CDR Global<sup>c</sup>, N**<br>(%) 0 | 189<br>(97.4) | 52<br>(100) | 71<br>(94.7) | 66<br>(98.5) | $p = 0.22$ | NA |
| **Clinical characteristics (adjusted for age, sex, and years of education)** |  |  |  |  |  |  |
| **GDS<sup>b</sup>,**<br>*mean ± SD* | 4.8 ±<br>4.8 | 4.8 ±<br>5.4 | 4.6 ±<br>4.1 | 5.0 ±<br>5.0 | $p = 0.90$ | NA |
| **GAD-7<sup>b</sup>,**<br>*mean ± SD* | 2.1 ±<br>3.1 | 2.3 ±<br>3.3 | 2.3 ±<br>3.5 | 1.7 ±<br>2.2 | $p = 0.27$ | NA |
| **WHOQOL-OLD<sup>b</sup>,**<br>*mean ± SD* | 94.6 ±<br>11.8 | 93.5 ±<br>13.2 | 92.1 ±<br>10.8 | 98.0 ±<br>11.2 | $p = 0.01$ | **SA < White, p = 0.04**<br>**Chinese < White, p = 0.005**<br>Chinese vs. SA, p = 0.57 |
| **Glucose levels**<br>**(HbA1c)<sup>b</sup> mean ±**<br>**SD (%)** | 5.8 ±<br>0.5 | 5.8 ±<br>0.5 | 6.0 ±<br>0.5 | 5.5 ±<br>0.3 | $p < 0.001$ | **SA > White, p < 0.001**<br>**Chinese > White, p < 0.001**<br>SA < Chinese, p = 0.11 |
| **Total**<br>**cholesterol<sup>d</sup> mean**<br>**± SD (mmol/L)** | 5.2 ±<br>1.2 | 5.0 ±<br>1.3 | 5.1 ±<br>1.3 | 5.3 ±<br>1.1 | $p = 0.46$ | NA |
| **HDL cholesterol<sup>d</sup>**<br>**mean ± SD**<br>**(mmol/L)** | 1.7 ±<br>0.5 | 1.6 ±<br>0.4 | 1.8 ±<br>0.5 | 1.8 ±<br>0.5 | $p = 0.003$ | **SA < White, p = 0.002**<br>Chinese vs White, p = 0.99<br>**SA < Chinese, p = 0.002** |
| **LDL cholesterol<sup>d</sup>**<br>**mean ± SD**<br>**(mmol/L)** | 2.9 ±<br>1.0 | 2.9 ±<br>1.1 | 2.8 ±<br>1.1 | 3.1 ±<br>0.9 | $p = 0.51$ | NA |
| **Non-HDL**<br>**cholesterol<sup>d</sup> mean**<br>**± SD (mmol/L)** | 3.5 ±<br>1.1 | 3.5 ±<br>1.2 | 3.4 ±<br>1.2 | 3.5 ±<br>1.0 | $p = 0.69$ | NA |
| **Triglycerides<sup>d</sup>**<br>**mean ± SD**<br>**(mmol/L)** | 1.1 ±<br>0.6 | 1.4 ±<br>0.7 | 1.1 ±<br>0.6 | 1.0 ±<br>0.5 | $p = 0.005$ | **SA > White, p = 0.001**<br>Chinese vs. White, p = 0.41<br>**SA > Chinese, p = 0.01** |
| **Systolic BP<sup>b</sup>**<br>**mean ± SD** | 131.7 ±<br>16 | 133.4 ±<br>13.2 | 131.9 ±<br>16.3 | 130.2 ±<br>17.5 | $p = 0.64$ | NA |
| **Diastolic BP<sup>b</sup>**<br>**mean ± SD** | 73.9 ±<br>9.0 | 72.5 ±<br>8.9 | 74.8 ±<br>9.2 | 73.8 ±<br>8.8 | $p = 0.63$ | NA |
| **Waist-to-hip**<br>**ratio<sup>e</sup>**<br>**mean ± SD** | 0.9 ±<br>0.1 | 0.9 ±<br>0.1 | 0.9 ±<br>0.1 | 0.9 ±<br>0.1 | $p = 0.51$ | NA |
| **BMI<sup>f</sup> mean ± SD**<br>**(kg/m<sup>2</sup>)** | 25.17 ±<br>4.23 | 25.8 ±<br>3.9 | 23.8 ±<br>3.8 | 26.2 ±<br>4.6 | $p = 0.001$ | SA vs. White, p = 0.66<br>**Chinese < White, p < 0.001**<br>**Chinese < SA, p = 0.006** |

**Abbreviations:** BMI: body mass index; BP: blood pressure; CDR: Clinical Dementia Rating; CFI: Cognitive Function Index; GAD-7: Geriatric Anxiety Scale-7; GDS: Geriatric Depression Scale; HbA1c: Hemoglobin A1c; HDL: High-density lipoprotein; LDL: Low-density lipoprotein; MoCA: Montreal Cognitive Assessment; NA: Not Applicable; SA: South Asian; SD: standard deviation; WHOQOL-OLD: World Health Organization Quality of Life – Older Adults Module
<sup>a</sup>1 missing value; <sup>b</sup>2 missing values; <sup>c</sup>6 missing values; <sup>d</sup>3 missing values; <sup>e</sup>4 missing values; <sup>f</sup>5 missing
values
\*Reported for participants who immigrated to Canada only (n=135)
Demographic characteristics report *P*-values representing results of ANOVA, Kruskal-Wallis, Chi-squared, or Fisher's Exact tests comparing groups.
Cognitive and clinical characteristics report *P*-values representing results of linear regression models controlling for age, sex, and years of education.
Post-hoc results are reported for contrasts where $p < 0.05$ .

With respect to follow-up, 99 participants completed Year 2 remote questionnaires (33 South Asian, 34 Chinese, and 32 NHW participants), with no dropouts. In addition, 46 have completed Year 3 in-person follow-up assessments (17 South Asian, 16 Chinese, and 13 NHW participants), with only 4 dropouts (3 Chinese, 1 NHW).

As shown in **Table 2**, age, sex, years of education, and annual income did not significantly differ across South Asian, Chinese, and NHW participants. As expected, South Asian and Chinese participants were less likely to be born in Canada, and more likely to report English as a second language compared to NHW participants. Compared to Chinese participants, South Asian participants were younger at the time of immigration and had a longer duration of residence in Canada.

### 3.2 Clinical Characteristics

Differences in clinical characteristics across ethnoracial groups were examined using linear regression models adjusted for age, sex, and years of education (**Table 2**). South Asian participants had significantly lower high-density lipoprotein (HDL) cholesterol and higher triglyceride levels than both Chinese and NHW participants. Both South Asian and Chinese participants had higher HbA1c levels compared to NHW participants. In contrast, Chinese participants had significantly lower body mass index (BMI) than both South Asian and NHW participants. South Asian and Chinese participants endorsed lower quality of life than NHW participants. No significant group differences were observed in depressive or anxiety symptoms, cholesterol levels (total, low-density lipoprotein, non-HDL), blood pressure, or waist-to-hip ratio.

### 3.3 Neuroimaging Data

Neuroimaging analyses focused on total grey matter volume (GMV) derived from FreeSurfer version 8.0 (see Supplementary Methods for preprocessing details). After adjustment for age, sex, years of education, and estimated total intracranial volume (eTIV), linear regression models indicated that both South Asian (β=−0.21, p = 0.008) and Chinese (β=−0.19, p=0.003) participants exhibited lower GMV relative to NHW participants, while no differences were observed between South Asian and Chinese participants (β=−0.02, p=0.80).

To explore potential explanations for these differences, we examined whether cardiometabolic factors mediated the association between ethnoracial group and GMV. Mediation analyses were conducted in R using the *mediation* package and focused on cardiometabolic variables that differed between groups, with models adjusted for age, sex, years of education, and eTIV. HbA1c significantly mediated the association between ethnoracial group and GMV. Specifically, HbA1c mediated GMV differences for South Asian versus NHW participants (average causal mediation effect [ACME]=−0.04, p=0.02) and for Chinese versus NHW participants (ACME=−0.06, p=0.008), accounting for approximately 21% and 34% of the total effect, respectively. In contrast, HDL cholesterol, triglycerides, and BMI did not mediate the relationship between ethnoracial group and GMV (**Supplementary Table 3**).

### 3.4 Cognitive and Eye Tracking Data

Cognitive (**Table 2**) and eye tracking (**Supplementary Table 4**) outcomes were examined across ethnoracial groups using linear regression models adjusted for age, sex, and years of education. No group differences were observed in global CDR scores. On the Montreal Cognitive Assessment (MoCA), a screening measure influenced by language and cultural background,^61^ both South Asian (vs. NHW: β=-0.73, *p*<0.001) and Chinese (vs. NHW: β=-0.66, *p*<0.001) participants scored significantly lower than NHW participants. Scores did not differ between the two Asian groups (South Asian vs. Chinese: β=-0.07, *p*=0.66). Results were unchanged after excluding participants with a global CDR of 0.5. Lower performance among South Asian and Chinese participants was most evident on MoCA items with greater English-language demands and reliance on Western cultural knowledge, including sentence repetition, verbal fluency, and animal naming. Correct responses for sentence repetition were observed in 74% of NHW participants, 29% of South Asian participants, and 9% of Chinese participants, with similar patterns for letter fluency (87% NHW, 81% South Asian, 70% Chinese) and animal naming (93% NHW, 79% South Asian, 80% Chinese)

The Cognitive Function Index (CFI) has been shown to be measurement invariant across race and ethnicity,^62^ indicating that it assesses subjective cognitive concerns equivalently across ethnoracial groups. On the CFI, both South Asian (vs. NHW: β=0.37, *p*=0.04) and Chinese (vs. NHW: β=0.66, *p*<0.001) participants endorsed significantly greater cognitive concerns than NHW participants, with no difference observed between the two Asian groups (South Asian vs. Chinese: β=-0.29, *p*=0.10). Results were unchanged after excluding individuals with a global CDR score of 0.5.

No significant group differences were observed on the eye-tracking measures (**Supplementary Table 4**), including IPAST pro-saccade reaction time, IPAST anti-saccade reaction time, IPAST direction errors, and VPCT novelty preference scores.

## 4.0 Discussion

Individuals of South Asian and Chinese descent are the largest and fastest-growing ethnoracial groups in Canada,^12^ yet they are underrepresented in ADRD research.^7–9,63^ The CAMERA study was designed to address this gap using a deep phenotyping approach, enabling examination of early biological, clinical, and lifestyle factors that may influence ADRD risk and resilience.

In the current sample, demographic profiles were broadly comparable across South Asian, Chinese, and NHW participants, with no group differences observed in age, sex, or years of education. Overall educational attainment was high (mean ∼16 years), aligning with patterns reported in other community-based aging and ADRD studies.^64,65^ This likely reflects both recruitment of research-engaged individuals and features of certain Canadian immigration pathways that prioritize higher educational attainment (e.g., points-based selection systems).^12^

Consistent with prior literature,^18,66^ cardiometabolic risk profiles differed across groups. South Asian participants exhibited a less favourable risk profile than NHW participants, including significantly lower HDL cholesterol, higher HbAc1 levels, and higher triglyceride levels. In contrast, and in line with some,^67^ but not all previous studies,^18^ Chinese participants exhibited higher HbAc1 levels than NHW participants, despite having significantly lower BMI. This pattern aligns with evidence that Asian populations, including Chinese individuals, may develop insulin resistance at lower BMI thresholds than NHW individuals.^68^

We further observed that both South Asian and Chinese participants had lower total GMV than NHW participants, with these differences mediated by HbA1c levels. This finding is consistent with prior cohort studies linking elevated HbA1c to markers of neurodegeneration.^69^ Together, these results suggest that glycemic dysregulation may be an important pathway contributing to brain atrophy in South Asian and Chinese participants.

South Asian and Chinese participants endorsed significantly greater cognitive concerns on the CFI compared to NHW participants, even when analyses were restricted to clinically normal individuals. Prior work has demonstrated that the CFI is measurement invariant across racial and ethnic groups, making it unlikely that these differences reflect measurement bias.^62^ This pattern is consistent with earlier studies in cognitively unimpaired adults reporting higher CFI scores among Asian participants relative to NHW peers.^70^ The reasons underlying greater endorsement of concerns among Asian participants will be examined in future analyses.

South Asian and Chinese participants scored lower on the MoCA than NHW participants. These differences likely reflect measurement-related factors rather than true differences in underlying cognitive ability. The MoCA has not been formally demonstrated to be measurement invariant in English-speaking Asian populations, and in the present study, group differences were primarily driven by items with greater reliance on English proficiency and Western cultural knowledge, including sentence repetition, letter fluency, and animal naming. Consistent with this interpretation, prior work has shown that standard MoCA cutoffs can overestimate cognitive impairment in ethnoracially diverse adults.^71^ Together, these findings suggest that MoCA performance may not accurately capture cognitive function in diverse samples and should be interpreted within the context of language and cultural background, highlighting the need for assessment tools that reduce linguistic and cultural bias.

These concerns motivated our inclusion of eye-tracking measures, which can be designed to reduce language-dependent and culturally biased stimuli and do not require verbal responses. Although we observed no group differences on eye-tracking measures, formal evaluation of measurement invariance following previously established procedures^72^ is planned to confirm that these tasks operate equivalently across ethnoracial groups.

A defining strength of CAMERA is its partnership with community stakeholders, which enhances the relevance, contextualization, and responsible interpretation of findings within the communities represented. Ongoing engagement with the Community Advisory Board (CAB) helps situate findings within participants’ lived experiences and supports communication of conclusions in ways that are meaningful and respectful to the communities involved. Participants frequently describe their involvement in CAMERA as collaborative, which may contribute to recruitment (including word-of-mouth referrals) and sustained engagement. Many participants have also expressed appreciation for receiving individualized results and for ongoing communication through quarterly newsletters and timely responses to questions. Together, these practices promote trust and ongoing engagement and are associated with high levels of participant-driven referrals.

To our knowledge, CAMERA is the only deep phenotyping study in North America designed specifically to examine ADRD risk and resilience in both South Asian and Chinese communities. The Asian Cohort for Alzheimer’s Disease (ACAD)^13^ represents one of the most closely related initiatives, as it includes Canadian participants in addition to those in the U.S., but differs from CAMERA in several key respects. ACAD focuses on individuals of Chinese, Korean, and Vietnamese ancestry and aims to characterize genetic and lifestyle factors primarily through self-report questionnaires within a large cohort of ∼5,000 participants. A key strength of ACAD is the use of translated cognitive assessments. In contrast, CAMERA emphasizes more intensive, multimodal deep phenotyping in a smaller cohort, incorporating neuroimaging and objective health measures to enable a more comprehensive characterization of ADRD risk and resilience.

Other large U.S.-based cohort studies provide important complementary context. The Multi-Ethnic Study of Atherosclerosis (MESA)^73^ was originally designed to examine subclinical atherosclerosis in individuals of Chinese, African American, Hispanic, and NHW backgrounds, but not South Asian. More recently, the MESA-MIND ancillary study incorporated brain imaging and cognitive assessments, extending MESA’s relevance to research brain aging. In contrast, the Mediators of Atherosclerosis in South Asians Living in America (MASALA)^74^ study focuses on South Asian adults and has generated rich longitudinal data on cardiovascular and metabolic risk, but does not include neuroimaging, ADRD biomarkers, or dementia-focused cognitive assessments.

Despite its many strengths, the CAMERA study has several limitations. First, all assessments are conducted in English, which may limit generalizability to individuals who are not proficient in English. This decision reflects practical and resource-related considerations, including the availability of validated assessment tools and appropriately trained staff. As a result, individuals with limited or no English proficiency may be underrepresented, and findings should be interpreted within this context. Although this approach does not capture the full linguistic diversity of these populations, it reflects aspects of the Canadian context, where some immigration pathways prioritize English (or French) proficiency (e.g., points-based selection systems). Second, although our neuropsychological test battery, including eye tracking tests, was designed to minimize language demands and culturally specific content, we recognize that even nonverbal cognitive tests can be influenced by cultural factors. It remains to be determined whether the eye-tracking measures included in CAMERA are invariant across the included ethnoracial groups, and this will be formally examined in future analyses. Finally, the exclusion of individuals with major cardiovascular or cerebrovascular events, along with the relatively high education and income levels of the sample, may further limit the generalizability of our findings. In addition, bilingualism and multilingualism in some participants may contribute to cognitive reserve and will be examined more explicitly in future analyses.

In sum, CAMERA represents an important step toward addressing the longstanding underrepresentation of South Asian and Chinese communities in ADRD research. By leveraging a deep-phenotyping approach, the study provides a platform for identifying potential risk and protective factors and clarifying their underlying mechanisms. As the cohort expands and longitudinal data accrue, CAMERA is well positioned to generate insights that may inform future large-scale studies and more tailored approaches to prevention and intervention for these communities.

## Data Availability

All data produced in the present study are available upon reasonable request to the authors.

## Acknowledgements

We would like to thank Ms. Aastha Vaidya for her contribution to the CAMERA study.

## Consent Statement

The CAMERA study was approved by the Research Ethics Board at Sunnybrook Health Sciences Centre (SHSC) in Toronto, Canada. All participants provided informed consent.

