## Supplementary Materials for "The Canadian multi-ethnic research on aging (CAMERA) study: Study design, participant characteristics, and preliminary findings"

### **Supplementary Table 1: Magnetic Resonance Imaging (MRI) Acquisition Parameters.**

| **Parameter** | **T1w-MPRAGE** | **T2w-SPACE** | **T2w-FLAIR** | **rsfMRI** | **dMRI** | **HCP** | **ASL** | **SWI** |
| --- | --- | --- | --- | --- | --- | --- | --- | --- |
| **TR (ms)** | 2400 | 3200 | 5000 | 800 | 5500 | 4800 | 4100 | 28.0 |
| **TE (ms)** | 2.28 | 408 | 388 | 37 | 99.20 | 106 | 36.76 | 20.0 |
| **TI (ms)** | 1060 | — | 1800 | — | — | — | — | — |
| **Echo Spacing (ms)** | 8.5 | 3.61 | 246 | 0.58 | 0.93 | 13.2 | 0.49 | — |
| **Flip Angle (°)** | 8 | — | — | 52 | 90 | 135 | 120 | 9 |
| **Bandwidth (Hz/Px)** | 210 | 723 | 751 | 2290 | 1488 | 130 | 2604 | 120 |
| **Field of View (mm)** | 256 | 240 | 256 | — | 208 | 150 | 240 | 240 |
| **EPI Factor** | — | — | — | 104 | 140 | — | 63 | — |
| **Voxel Size (mm)** | 0.8 × 0.8 × 0.8 | 0.9 × 0.9 × 0.9 | 1 × 1 × 1 | 2 × 2 × 2 | 1.6 × 1.6 × 1.6 | 0.4 × 0.4 × 2.0 | 2.5 × 2.5 × 2.5 | 0.6 × 0.6 × 3.0 |

**Structural MRI Pre-processing**

Structural MRI data were preprocessed following standard procedures. Raw images were first converted from DICOM to NIfTI format using **dcm2niix**.^1^ Images were then axis-aligned and centered using **nifti_align**.^2^. Cortical and subcortical reconstruction and volumetric segmentation were subsequently performed using the default **recon-all** pipeline in FreeSurfer v8, with both T1-weighted and T2-weighted images provided as input.

### **Supplementary Table 2:** CAMERA Questionnaires

| **Variable** | **Data Collected and Questionnaires Administered** |
| --- | --- |
| **Demographic information** | Age  Sex assigned at birth (male, female, intersex) Gender identity (man, woman, transgender man, transgender woman, non-binary, two-spirit)  Handedness  Sexual orientation  Race  Ethnicity  Marital status and number of children  Childhood and current household compositions  Caregiver status  Lifestyle factors (alcohol use and smoking)  Social network and loneliness |
| **Socioeconomic status** | Education (number of years, location, and language)  Self-reported household gross income  Location of residence (urban/rural)  Subjective rating of social status  Employment status  Occupation coded for prestige and cognitive complexity using the Standard Occupational Classification^3^ |
| **Immigration history** | Immigration status (country of origin, year entered Canada, and age at immigration)  Ancestral country of origin  Reason for immigration to Canada  Age of immigration  Year arrived in Canada  Location of arrival  Method of immigration  Presence of social and community ties in Canada |
| **Linguistic background** | Native language  Age learned English  Number of languages spoken |
| **Acculturation** | Vancouver Index of Acculturation^4^  Acculturation Questionnaire |
| **Environmental and community factors** | Modified Environmental Profile of a Community's Health (EPOCH)^5^ |
| **Medical history** | Past and current chronic medical conditions  Family health history of chronic medical conditions  Current medications  Birth history  Learning/early attention difficulties  Health status |
| **Mental health and personality characteristics** | Geriatric Depression Scale (Long Form) (GDS-30)^6^  Generalized Anxiety Disorder Scale (GAD-7)  Behavioural Inhibition System (BIS) / Behavioural Activation System (BAS) |
| **Physical activity and Sedentary behavior** | International Physical Activity Questionnaire (IPAQ)^7^  International Sedentary Assessment Tool (ISAT)^8^ |
| **Sleep Quality** | Pittsburgh Sleep Quality Index (PSQI)^9^ |
| **Quality of life** | World Health Organization Quality of Life Scale – Older Adults (WHOQOL-OLD)^10^ |
| **Perceived discrimination** | Perceived Discrimination Scale (PDS)^11^ |
| **Cognitive concerns** | Cognitive Function Index (CFI) and CFI-Study Partner^12,13^ |
| **Functional status** | Functional Activities Questionnaire (FAQ) (informant-reported)^14^ |
| **Sex-Specific history** | Female-specific health questions: Pregnancy history, menopausal history, menopausal symptoms, and current and past use of hormone therapy and hormonal contraceptives  Male-specific health questions: Testosterone replacement therapy use, anabolic steroids use, and hormone-related conditions and treatments |
| All questionnaires are administered annually via REDCap. | |

### **Supplementary Table 3**: Mediation models examining cardiometabolic contributors to ethnoracial differences in grey matter volume.

| **Mediator** | **South Asian and NHW** | **Chinese and NHW** |
| --- | --- | --- |
| **HbA1c**^a^ | ACME = -0.04; *p* = 0.02  ADE = -0.16; *p* = 0.05  Total effect t = -0.21; *p* = 0.02 | ACME = -0.06; *p*= 0.008  ADE = -0.13; *p* = 0.06  Total effect = -0.19; *p* = 0.004 |
| **HDL cholesterol**^b^ | ACME = -0.02; *p* = 0.26  ADE = -0.19; *p* = 0.03  Total effect = -0.21; *p* = 0.01 | ACME = 0.0008; *p* = 0.87  ADE=-0.19; *p*=0.006  Total effect=-0.19; *p*=0.004 |
| **Triglycerides**^b^ | ACME=-0.02; *p*=0.34  ADE=-0.19; *p*=0.04  Total effect=-0.21; *p*=0.01 | ACME=-0.004; *p*=0.66  ADE=-0.19; *p*=0.02  Total effect=-0.19; *p*=0.008 |
| **BMI**^b^ | ACME=0.003; *p*=0.79  ADE=-0.20; *p*=0.01  Total effect=-0.19; *p*=0.01 | ACME=0.03; *p*=0.11  ADE=-0.20; *p*=0.004  Total effect=-0.18; *p*=0.01 |
| Abbreviations: ACME: average causal mediation effect; BMI: body mass index; GMV: grey matter volume; eTIV: estimated total intracranial volume; HbA1c: Hemoglobin A1c; HDL: High-density lipoprotein; NHW: non-Hispanic White  ^a^11 missing participants (n=189)  ^b^12 missing participants (n=188)  ^c^14 missing participants (n=186)  Reported estimates are standardized estimates. | | |

### **Supplementary Table 4:** Ethnoracial differences in eye-tracking measures from adjusted linear regression models.

| **Comparison** | **IPAST PRO SRT^a^** | **IPAST ANTI SRT^a^** | **IPAST ANTI Direction Error Rate^a^** | **VPCT Saccade Novelty Score^a^** |
| --- | --- | --- | --- | --- |
| **South Asian vs. White**^a^ | β=0.05;  *p*=0.79 | *β*=0.02;  *p*=0.93 | *β*=-0.34;  *p*=0.08 | *β*=0.18;  *p*=0.36 |
| **Chinese vs. White**^a^ | *β*=0.27;  *p*=0.14 | *β*=0.15;  *p*=0.42 | *β*=-0.29;  *p*=0.10 | *β*=-0.05;  *p*=0.78 |
| **South Asian vs. Chinese**^a^ | *β*=-0.21;  *p*=0.29 | *β*=-0.13;  *p*=0.51 | *β*=-0.05;  *p*=0.81 | *β*=0.23;  *p*=0.25 |
| Abbreviations: IPAST: interleaved Pro/Anti-Saccade Task; SRT: saccadic reaction time; VPCT: Visual Paired Comparison Task  ^a^32 missing participants: 23 did not complete eye tracking, and 9 did not have valid data.  Reported estimates are standardized beta (β) coefficients from linear regression models. | | | | |
